# Characteristics and Outcomes of COVID-19 Patients Presumed to be Treated with Sotrovimab in NHS Hospitals in England

**DOI:** 10.1101/2023.02.08.23285654

**Authors:** Vishal Patel, Bethany Levick, Stephen Boult, Daniel C. Gibbons, Myriam Drysdale, Emily J. Lloyd, Moushmi Singh, Helen J. Birch

## Abstract

**Introduction:** There is limited real-world evidence describing the effectiveness of early treatments for Coronavirus disease 2019 (COVID-19) during the period where Omicron was the dominant variant. Here we describe characteristics and acute clinical outcomes in patients with COVID-19 treated with a monoclonal antibody (mAb; presumed to be sotrovimab) across six distinct periods covering the emergence and subsequent dominance of Omicron subvariants (BA.1, BA.2 and BA.5) in England.

**Methods:** Retrospective cohort study using data from Hospital Episode Statistics database between 1^st^ January – 31^st^ July 2022. Included patients were aged ≥12 years and received a mAb delivered by a National Health Service (NHS) hospital as a day-case, for which the primary diagnosis was COVID-19. Patients were presumed to have received sotrovimab on the basis of available NHS data showing that 99.98% of individuals who received COVID-19 treatment during the period covered by the study were actually treated with sotrovimab. COVID-19-attributable hospitalisations were reported overall and across six distinct periods of Omicron sub-variant prevalence. A multivariate Poisson regression model was used to estimate incidence rate ratios for each period. Subgroup analyses were conducted in patients with severe renal disease and active cancer.

**Results:** In total, 10,096 patients were included. The most common high-risk comorbidities were Immune-Mediated Inflammatory Disorders (43.0%; *n* = 4,337), severe renal disease (14.1%; *n* = 1,422), rare neurological conditions (10.4%; *n* = 1,053) and active cancer (9.0%; *n* = 910). The proportions of patients with a COVID-19-attributable hospitalisation was 1.0% (*n* = 96), or with a hospital visit due to any cause was 4.6% (*n* = 465) during the acute period. The percentage of patients who died due to any cause during the acute study period was 0.3% (*n* = 27). COVID-19-attributable hospitalisation rates were consistent among subgroups and no significant differences (p-values ranged from 0.13 to 0.64) were observed across periods of Omicron subvariants.

**Conclusion:** Low levels of COVID-19-attributable hospitalisations and deaths were recorded in mAb-treated patients. Results were consistent for patients with severe renal disease and active cancer. No evidence of differences in hospitalisation rates were observed whilst Omicron BA.1, and BA.2 or BA.5 subvariants were predominant, despite reported reductions in in vitro neutralisation activity of sotrovimab against BA.2 and BA.5.

## INTRODUCTION

Coronavirus disease 2019 (COVID-19), which is caused by infection with the severe acute respiratory syndrome coronavirus 2 (SARS-CoV-2), has been associated with a substantial clinical and economic burden worldwide [1, 2].

Sotrovimab is a dual-action Fc-engineered human monoclonal antibody (mAb) that was developed for the treatment of COVID-19 and targets a conserved epitope in the SARS-CoV-2 spike protein distinct from the angiotensin-converting enzyme-2 (ACE2) receptor binding motif [3]. The phase 2/3 COVID-19 Monoclonal antibody Efficacy Trial-Intent to Care Early (COMET-ICE; NCT04545060) trial assessed the efficacy and safety of sotrovimab administered intravenously in high-risk patients with mild-to-moderate COVID-19 and was conducted during a period of wildtype SARS-CoV-2 predominance. Final results for the primary endpoint showed a 79% (95% confidence interval: [50%, 91%], *P* < 0.001) relative risk reduction in all-cause >24-hour hospitalisation or death with sotrovimab treatment compared with placebo [4].

Sotrovimab received approval from the European Medicines Authority [5] and Medicines & Healthcare products Regulatory Agency in December 2021 [6] for the ambulatory treatment of mild-to-moderate COVID-19 in adults and adolescents (≥12 years of age) who are at increased risk of developing severe disease. Sotrovimab is administered as a day-case intravenous infusion where the patient is admitted electively with the intention of not using a hospital bed overnight [6]. In addition to sotrovimab, high-risk COVID-19 patients in the UK can also be eligible for early treatment with nirmatrelvir/ritonavir, molnupiravir or remdesivir [7, 8].

In vitro pseudotyped viral assays have assessed neutralisation activity of sotrovimab against Omicron variants, with 2.7-, 16- and 22.6-fold changes in IC50 versus wild-type reported for BA.1, BA.2 and BA.5, respectively [3, 9]. The clinical implications of reduced in vitro neutralisation are unknown and there is no validated clinical pharmacology model for sotrovimab that can reliably predict clinical efficacy from in vitro neutralisation.

Here we describe the real-world use of, and clinical outcomes in a population with assumed treatment with sotrovimab for the management of high-risk patients with COVID-19 in National Health Service (NHS) hospitals across England, at times when Omicron BA.1, BA.2 and BA.5 were predominant.

## METHODS

### Study Design and Data Source

This was a retrospective cohort study using data from the Hospital Episode Statistics (HES) database. HES is a data warehouse containing records of hospital diagnoses, procedures, treatments, health care resource utilisation (HCRU) and associated costs for all patients admitted to NHS hospitals in England.

### Identification of Presumed Sotrovimab Administration

HES describes diagnoses and procedures associated with episodes of care without direct reporting of pharmacy data. Whilst we were unable to directly ascertain sotrovimab administration, weekly data for individuals receiving COVID-19 treatments showed that, during the study period, the vast majority of non-hospitalised patients being treated with a mAb were actually treated with sotrovimab (30,234 patients out of a total of 30,241 [99.98%] – as per the report published on 5^th^ January 2023) [10]. As such, episodes identified as day-case admissions that were associated with both a primary diagnosis of COVID-19 (ICD-10 U07.1) [11] and a record of intravenous mAb administration (OPCS-4 X89.2, per NHS Digital guidance) [12] were deemed to represent sotrovimab administration for the purposes of this study. ICD-10 U07.2 code, which translates to “COVID-19, virus not identified” as per the World Health Organisation IDC-10 2019 guidance, was used to confirm absence of COVID-19 diagnosis [11].

### Population

To be eligible for inclusion, patients had to have a record of mAb administration (OPCS-4 X89.2) within a spell occurring between 1^st^ of January 2022 and 31^st^ of July 2022 that was identified as a day-case admission and was associated with a primary diagnosis of COVID-19 (ICD-10 U07.1) in HES. The start date of the earliest qualifying spell was considered a patient’s index date and the spell was considered the index spell. To be included in analyses, patients had to be aged ≥12 years as of their index date.

Patients whose index spell had a length of stay greater than one day or had another record associated with a mAb administration in the 28 days prior to index (or following the first event other than where given as part of inpatient care in the study period), were excluded from the study.

The baseline period, during which comorbidities were identified, was defined as the 365 days prior to the index date. A patient’s acute period, during which outcomes were evaluated, was defined as the 28 days following the index date.

### Study Periods

The study was divided into six distinct periods that reflected the dynamics of Omicron BA.1, BA.2 and BA.5 subvariant activity (Table 1). These periods were defined based on the prevalence of SARS-CoV-2 infections with these variants using sequencing data that were reported in the weekly technical briefing reports published by the UK Health Security Agency (HAS) [13].

**Table 1.** Study periods.

|  | **Omicron subvariants prevalence [13]** | **Duration** |
| --- | --- | --- |
| Period 1 | BA.2 prevalence <25%<br>(predominant variant: BA.1) | 1 January 2022 – 6 February 2022 |
| Period 2 | BA.2 prevalence ≥25% but <75% | 7 February 2022 – 27 February 2022 |
| Period 3 | BA.2 prevalence ≥75%, | 28 February 2022 – 1 May 2022 |
| Period 4<sup>a</sup> | BA.5 prevalence <25% | 2 May 2022 – 31 May 2022 |
| Period 5 | BA.5 prevalence ≥25% but <75% | 1 June 2022 – 3 July 2022 |
| Period 6 | BA.5 prevalence ≥75% | 4 July 2022 – 31 July 2022 |

<sup>a</sup>Starting with period 4, a declining prevalence of Omicron BA.2 and increasing prevalence of Omicron BA.5 was observed; the main circulating variants were Omicron BA.4 and BA.5.

### Patient Characteristics and Study Outcomes

Patient characteristics (age, gender, ethnicity, presence of specific comorbidities that indicate a high risk of developing severe COVID-19, and previous admissions for COVID-19), collated from already available baseline data or information captured during index spell, were described for the overall cohort.

The primary outcomes of this study – COVID-19 attributable hospitalisations and all-cause hospitalisations and deaths – were captured during the 28-day post-index acute period. A COVID-19-attributable hospitalisation was defined as a hospital visit in which COVID-19 was listed in the primary diagnosis field during the acute period. All-cause hospitalisations were defined as any hospital visits which occurred during the acute period. Deaths reported in the acute period were also reported.

The secondary outcomes of this study described the proportion of patients treated during each of the six distinct 3-to 8-week periods of Omicron subvariants activity; the occurrence of COVID-19 attributable hospitalisations during the acute period was described for each of the six periods, and treatment outcomes were compared between period 1 and the other five periods.

Primary outcomes were also reported in two sub-populations of interest: severe renal disease or active cancer. Severe renal disease (based on ICD-10 diagnosis codes) included patients with chronic kidney disease stage 4 or 5, those in receipt of peritoneal dialysis or haemodialysis or those with a kidney transplant. Active cancer was defined as people with cancer (based on a relevant cancer code at any time prior to assumed sotrovimab administration) who had received chemotherapy or radiotherapy within the 12 months prior to their index date.

### Data Analysis

Continuous variables, such as age, were summarised using mean, standard deviation, median, interquartile range and range. Categorical variables, such as gender, were described using frequencies and percentages. Small number suppression was applied for all small numbers up to and including 7 by being rounded to the nearest 5 (regardless of the actual number). These values were replaced with an asterisk.

Incidence rates (per 100 patient-days) within 28 days were calculated as the number of hospitalisations divided by the total person time observed in days and amplified by 100. To compare incidence rates between period 1 (Omicron BA.1 predominance and BA.2 prevalence of less than 25%), and each of the other five periods, a multivariate Poisson regression model was used to estimate incidence rate ratios and associated confidence intervals for each period. The estimates were adjusted for patient age, previous COVID-19 admission, the presence of evidence of at least one high-risk comorbidity in the patient record and time period of index.

## RESULTS

### Patient Demographics and Baseline Characteristics

In total, 10,096 patients were eligible for inclusion in the study (Table 2). The mean age of patients was 56.4 years and 42.0% of the study population was female (*n* = 4,238). The percentage of patients who had a previous hospital admission in which COVID-19 was listed as a primary or underlying diagnosis was 3.0% (*n* = 298). Of the high-risk comorbidities, the most frequently reported were Immune-Mediated Inflammatory Disorders (IMID) (43.0%, *n* = 4,337), severe renal disease (14.1%, *n* = 1,422), rare neurological conditions (10.4%, *n* = 1,053) and active cancer (9.0%, *n* = 910). There was no evidence of presence of high-risk comorbidities (based on available diagnosis codes) in 26.1% (*n* = 2,633) of included patients.

**Table 2.** Patient characteristics.

|  | **Patients**<br>**(n = 10,096)** |
| --- | --- |
| **Age, years** |  |
| Mean (SD) | 56.40 (16.4) |
| Median (IQR) | 57 (44–69) |
| **Age group, years, n (%)** |  |
| 12–54 | 4,412 (43.7) |
| 55–64 | 2,278 (22.6) |
| 65–74 | 1,961 (19.4) |
| ≥75 | 1,445 (14.3) |
| **Female sex, n (%)** | 4,238 (42) |
| **Ethnicity, n (%)<sup>a</sup>** |  |
| White | 6,955 (68.9) |
| Asian/Asian British | 619 (6.1) |
| Black/Black British/Caribbean or African | 239 (2.4) |
| Mixed | 114 (1.1) |
| Other | 329 (3.3) |
| Unknown | 1,840 (18.2) |
| **Previous admission for COVID-19, n (%)** | 298 (3.0) |
| **High-risk comorbidities, n (%)** |  |
| Active cancer | 910 (9.0) |
| Down syndrome | 107 (1.1) |
| HIV | <sup>b</sup> |
| Immune deficiencies | 338 (3.3) |
| Patients being treated for immune-mediated inflammatory disorders | 4,337 (43.0) |
| Patients with haematological diseases and stem cell transplant recipients | 602 (6.0) |
| Patients with liver disease | 438 (4.3) |
| Rare neurological conditions | 1,053 (10.4) |
| Severe renal disease | 1,422 (14.1) |
| Solid organ transplant recipients | 280 (2.8) |
| **No comorbidity, n (%)** | 2,633 (26.1) |

COVID-19, Coronavirus disease 2019; IQR, interquartile range; SD, standard deviation.
<sup>a</sup>Percentages calculated based on removal off ‘unknown’ group from the denominator; <sup>b</sup>Small number suppression applied.

### Acute Period Outcomes

Acute period outcomes during the full study period (1^st^ January 2022 – 31st July 2022) are presented in Table 3. COVID-19-attributable hospitalisations were recorded in 1.0% (*n* = 96) of patients. The percentage of patients who had a hospital visit due to any cause during the acute period following their sotrovimab treatment was 4.6% (*n* = 465). Overall, 0.3% (*n* = 27) of patients were recorded as having died due to any cause during the acute period.

**Table 3.** Overall acute period (28 days following index) outcomes.

|  | **Patients**<br>**(*n* = 10,096)** |
| --- | --- |
| COVID-19 attributable hospitalisation, n (%) | 96 (1) |
| Any cause hospitalisation, n (%) | 465 (4.6) |
| All-cause deaths, n (%) | 27 (0.3) |

*COVID-19*, Coronavirus disease 2019.

### Acute Period Outcomes for Patients with Advanced Renal Disease and Active Cancer

Among 1,422 patients with severe renal disease, 1.3% (*n* = 18) had a COVID-19-attributable hospitalisation during the acute period, 6.8% (*n* = 97) had a non-elective hospitalisation for any cause and 0.3% (*n* = 4) died due to any cause (Table 4).

**Table 4.** Acute period outcomes (28 days following index) among patients with severe renal disease and active cancer.

|  | **Severe renal disease**<br>**(*n* = 1,422)** | **Active cancer**<br>**(*n* = 910)** |
| --- | --- | --- |
| COVID-19 attributable hospitalisation, *n* (%) | 18 (1.3) | 10 (1.1) |
| Any cause hospitalisation, *n* (%) | 97 (6.8) | 89 (9.8) |
| All-cause deaths, *n* (%) | 4 (0.3) | 7 (0.8) |

COVID-19, Coronavirus disease 2019.

Out of 910 patients who were identified as having an active cancer, 1.1% (*n* = 10) had a COVID-19-attributable hospitalisation during the acute period, 9.8% (*n* = 89) had a hospitalisation for any cause and 0.8% (*n* = 7) died due to any cause (Table 4).

### Acute Period Outcomes Across Periods of Omicron Subvariants Prevalence

Acute period outcomes according to the time of diagnosis in the six periods of Omicron subvariants prevalence are shown in Table 5. The proportions of patients with a COVID-19-attributable hospitalisation across periods 1 to 6 were 1.0% (*n* = 22/2,102), 1.3% (*n* = 13/993), 1.0% (*n* = 37/3,884), 1.0% (*n* = 6/573), 1.4% (*n* = 16/1,161) and 0.7% (*n* = 10/1,383), respectively. This equated to an incidence rate per 100 patient-days of 0.040 for period 1, 0.050 for period 2, 0.036 for period 3, 0.040 for period 4, 0.052 for period 5 and 0.028 for period 6. A multivariate Poisson regression model found no evidence of significant differences in incidence of COVID-19 hospitalisations for periods 2–6 (p values ranged from 0.13 to 0.83) relative to period 1 adjusted for age at diagnosis, previous admission for COVID-19 or evidence of at least one high risk condition (Table 5).

**Table 5.** Acute period outcomes (28 days following index) across periods of Omicron subvariants prevalence.

|  | **Period 1**<br>**(BA.1**<br>**predominant,**<br>**BA.2 < 25%**<br>**prevalence)**<br>**(n = 2 102)** | **Period 2**<br>**(25% > BA.2**<br>**< 75%**<br>**prevalence)**<br>**(n = 993)** | **Period 3**<br>**(BA.2 > 75%**<br>**prevalence)**<br>**(n = 3 884)** | **Period 4**<br>**(BA.5 < 25%**<br>**prevalence)**<br>**(n = 573)** | **Period 5**<br>**(25% > BA.5**<br>**< 75%**<br>**prevalence)**<br>**(n = 1 161)** | **Period 6**<br>**(BA.5 > 75%**<br>**prevalence)**<br>**(n = 1 383)** |
| --- | --- | --- | --- | --- | --- | --- |
| COVID-19 attributable hospitalisation, n (%) | 22 (1.0) | 13 (1.3) | 37 (1.0) | 6 (1.0) | 16 (1.4) | 10 (0.7) |
| Incidence rate per 100 patient-days | 0.040 | 0.050 | 0.036 | 0.040 | 0.052 | 0.028 |
| Incidence rate ratio<sup>a</sup> (95% CI) | REF | 1.16<br>(0.58–2.31) | 0.76<br>(0.44–1.30) | 0.8<br>(0.32–1.99) | 1.07<br>(0.56–2.06) | 0.56<br>(0.26–1.19) |
| p-value | REF | 0.67 | 0.31 | 0.63 | 0.83 | 0.13 |

*CI*, confidence interval; *REF*, reference group.
<sup>a</sup>Incidence of hospitalisation = (hospitalisations observed/total person time in days) x 100.

## DISCUSSION

We investigated patient characteristics and outcomes (COVID-19-attributable hospitalisations and all-cause hospitalisations and deaths) in patients diagnosed with COVID-19 who received sotrovimab administered in NHS hospitals across England. The results of this study demonstrate that people who were treated with sotrovimab in England between 1^st^ January and 31^st^ July 2022 experienced low levels of COVID-19-attributable hospitalisations during the 28 days following treatment administration. COVID-19-attributable hospitalisations were also low in subgroups of people with severe kidney disease and active cancers. Continuous low rates of clinical outcomes such as all-case and COVID-19-attributable hospitalisations or deaths were reported across subvariant predominance periods (BA.1, BA.2 and BA.5). Moreover, the analysis of COVID-19-attributable hospitalisation rates between Omicron BA.1 (period 1), BA.2 (periods 2,3) and BA.5 activity (periods 4–6) indicated that there was no evidence of difference between period 1 and the other five periods.

The lack of pharmacy data in HES required indirect identification of assumed treatment with sotrovimab. Nonetheless, our results are consistent with those from a recent study, conducted between 16^th^ December 2021 and 10^th^ February 2022 using the OpenSAFELY-TPP platform, which reported that 0.96% of patients confirmed to have been treated with sotrovimab had a COVID-19-attributable hospitalisation or death within 28 days of treatment [14]; in our study, 1.0% of patients who were assumed to have been treated with sotrovimab experienced a COVID-19-attributable hospitalisation in the 28-day post-treatment acute period. The results are also similar to those of another recently completed analysis that used data from the Discover database in North-West London, which reported 0.7% of people confirmed to have been treated with sotrovimab experiencing a COVID-19-attributable hospitalisation during the 28 days following treatment (study period was 1^st^ December 2021 – 31^st^ May 2022 with subvariants predominance as follows: Omicron BA.1 from 1^st^ December 2021 – 28^th^ February 2022 and Omicron BA.2 from 1^st^ March 2022 – 31^st^ May 2022) [15]. Similarly, our findings of low rates of COVID-19-attributable deaths and hospitalisations in patients with advanced kidney disease are consistent with a those from a recent study in non-hospitalised patients with COVID-19 on kidney replacement therapy; treatment with sotrovimab resulted in a substantially lower risk of severe COVID-19 outcomes compared with molnupiravir during periods of Omicron BA.1 through to BA.5 subvariant dominance [16]. Finally, our findings are also consistent with the most recent data from the OpenSAFELY-TPP platform, comparing the effectiveness of sotrovimab and Paxlovid in preventing severe COVID-19 outcomes when different subvariants of Omicron were dominant [17]; the risk of severe outcomes was similar between the treatment groups, with no changes observed due to circulation of the BA.5 subvariant.

Recent studies have demonstrated that the Omicron BA.2 variant is similar in severity to the Omicron BA.1 variant [13, 14, 18, 19], although it may have increased severity in certain populations such as the elderly [20]. Our large population-based study across England contributes to the overall favorable weight of evidence to support clinical benefit of sotrovimab as an early treatment for COVID-19 through Omicron sub-variant predominance periods, especially for patients at higher risk of developing severe symptoms such as those with severe renal diseases and active cancer. Moreover, our findings also confirm those of a recent study that reported similar proportions of hospital admissions between sequence-confirmed Omicron BA.1 and BA.2 cases treated with sotrovimab [21]. In addition, our study further extends these findings by also assessing patients treated during periods of Omicron BA.5 prevalence.

These data, in conjunction with preclinical data supporting *in vitro* and *in vivo* antiviral activity of sotrovimab against Omicron BA.2 and Omicron BA.5 variants, reinforce the lack of validated models to predict correlates of efficacy based solely on *in vitro* neutralisation [22, 23]. The variability of *in vitro* results based on cell lines and assay systems and a lack of models to incorporate the role of Fc effector function (which triggers the body’s own innate immune cells to fight SARS-CoV-2 infection, thus contributing to sotrovimab effectiveness) may also compromise the ability to reproduce clinical effects *in vitro*. Therefore, the totality of available evidence including *in vitro, in vivo* and observational data should be considered when determining treatment options for early SARS-CoV-2 infections.

An important limitation of this study is the single-arm design, which prevented any comparisons with a reference group of patients. Administration of oral anti-virals is not captured within HES due to the lack of pharmacy data, therefore comparison with these agents (or confirmation of untreated groups) was not feasible with this data source. The absence of accurate data for SARS-CoV-2 positive infections in the community also contributed to the absence of an untreated comparator group.

Additionally, it is known that there is under-reporting of comorbidities within the HES database [24], and therefore the characterisation of high-risk comorbidities amongst sotrovimab-treated people may not be complete. In the current study, 26% of the total population did not have any comorbidities recorded. Moreover, recording only the comorbidities within a period of 12 months will also bring bias towards identification of clinically impactful and active comorbidities. Furthermore, the classification of each patient as a high-risk case relies on the associated diagnoses being recorded with an admission event for the identified patients. This may result in underestimation of some high-risk conditions, further compounded by the lack of pharmacy data on prescribed medicines. Also, as a given comorbidity has to have been severe enough to warrant review in hospital, and as many regular reviews of chronic conditions were likely deprioritised during the pandemic, this may have led to under-reporting. As a high-cost drug sotrovimab is unlikely to be approved for patients without a diagnosis fitting the eligibility criteria in the latest guidance [25]. Within this cohort, the variable as described could then more strictly be interpreted as acting as a proxy for those patients requiring recent hospital care where their diagnosis is noted.

Confirmatory polymerase chain reaction test results for COVID-19 were not available for the population of this study. However, initial studies suggest clinical coding of COVID-19 in HES is of good quality (England 2021), and an expanded COVID-19 clinical coding policy had been in place for over a year at the time of the study period, so any impacts are expected to be minimal [26, 27]. It is not possible to consistently distinguish planned and unplanned single overnight stays in HES data; therefore, in order to restrict included patients to the directed use of sotrovimab, all overnight stays were excluded. This may exclude some patients who are effectively hospitalised on day 0 following their treatment, although they would have to deteriorate substantially immediately after receiving their sotrovimab treatment and would probably not be eligible to receive sotrovimab in the NHS in England [6]. Lastly, COVID-19 vaccination status, which is likely to be linked to the probability of subsequently being admitted due to COVID-19, is also not available in the study dataset. However, vaccination rates in the study population are expected to be higher than in the general population due to their higher risk for poor COVID-19 outcomes and a longer time in which the vaccine was available to them.

## CONCLUSION

Patients assumed to have been treated with sotrovimab experienced low levels of COVID-19-attributable hospitalisations and all cause deaths across periods of different Omicron subvariant prevalence. The results were consistent within subgroups of patients with severe renal disease and active cancer, as well as across periods of Omicron BA.1, BA.2 and BA.5 subvariants activity. No evidence of differences in hospitalisation rates were observed during different periods aligned with prevalence of Omicron BA.1 and periods of BA.2 or BA.5 sub-variant predominance.

## Data Availability

The datasets generated during and/or analysed during the current study are available in the NHS-Digital via the Data Access Request Service (DARS) repository. Copyright ©2023 Re-used with the permission of NHS Digital.

## Medical Writing, Editorial and Other Assistance

Editorial support (in the form of writing assistance, including preparation of the draft manuscript under the direction and guidance of the authors, collating and incorporating authors’ comments for each draft, assembling tables and figures, grammatical editing and referencing) was provided by Mirela Panea of Luna, OPEN Health Communications, in accordance with Good Publication Practice (GPP) guidelines (www.ismpp.org/gpp-2022).

## Authorship

All named authors meet the International Committee of Medical Journal Editors (ICMJE) criteria for authorship for this manuscript, take responsibility for the integrity of the work as a whole and have given final approval for the version to be published.

## Author contributions

All authors contributed to the study conception and design. The study analysis was performed by Stephen Boult. Interpretation of the results was performed by Vishal Patel, Bethany Levick, Stephen Boult, Daniel C. Gibbons and Myriam Drysdale. All authors wrote the manuscript, interpreted the data and critically revised the manuscript. All authors read and approved the final manuscript and agree to be accountable for all aspects of the work.

## Compliance with ethical standards

This study complies with all applicable laws regarding subject privacy. Data were aggregated and counts less than five were suppressed in line with IG suppression rules. No direct subject contact or primary collection of individual human subject data occurred. Study results were in tabular form and aggregate analyses that omits subject identification, therefore informed consent, ethics committee or IRB approval are not required. Any publications and reports will not include subject identifiers.

Harvey Walsh Ltd (Part of the OPEN Health group) are licenced by NHS Digital to receive Hospital Episode Statistics data under data sharing agreement DARS-NIC-05934-M7V9K and follow NHS-Digital guidelines. The Independent Group Advising on the Release of Data (IGARD) advisory body to NHS Digital gave approval for the use of HES data.

## Availability of data and material

The datasets generated during and/or analysed during the current study are available in the NHS-Digital via the Data Access Request Service (DARS) repository. Copyright © 2023 Re-used with the permission of NHS Digital.

